# Acute Epstein Barr Virus is a risk factor for severe malaria in infants under 24 months

**DOI:** 10.1101/2025.04.29.25326585

**Authors:** Wayne T. Cheng, Balotin Fogang, Aarti Jain, D Huw Davies, Philip L Felgner, Carole Eboumbou, Paul N. Koki, Samuel H. Speck, Chester J. Joyner, Lawrence S Ayong, Tracey J Lamb

**Affiliations:** Department of Infectious Diseases, College of Veterinary Medicine, University of Georgia, Athens, GA, United States; Center for Tropical and Emerging Global Diseases, University of Georgia, Athens, GA, United States; Center for Vaccines and Immunology, University of Georgia, Athens, GA, United States; Molecular Parasitology Laboratory, Centre Pasteur du Cameroun, BP 1274 Yaounde, Cameroon; Vaccine Research and Development Center, Department of Physiology and Biophysics, University of California, Irvine, Irvine, CA, USA; Faculty of Medicine and Pharmaceutical Sciences, University of Douala, BP 2701 Douala, Cameroon; Mother and Child Centre of the Chantal Biya Foundation, Pediatric Service, Yaoundé, Cameroon; Department of Pediatrics, University of Yaoundé I, Cameroon; Department of Microbiology and Immunology, Emory University School of Medicine and the Emory Vaccine Center, Emory University, Atlanta, Georgia, USA; Department of Pathology, University of Utah, 15 N Medical Drive, Salt Lake City 84112, USA

**Keywords:** Co-infection, EBV, *P. falciparum*, humoral response, protein microarray, latency

## Abstract

**Background:** Primary Epstein Barr Virus (EBV) infection occurs during late adolescence and is characterized by the symptomatic manifestation of infectious mononucleosis (IM). Primary EBV infection in malaria-endemic areas often occurs in young children by the age of 2 and is generally asymptomatic. Acute EBV infection in children of this age results in humoral immune suppression to unrelated antigenic challenges for approximately 4 weeks. Whether acute EBV in infants similarly suppresses the development of antibody responses against *Plasmodium falciparum (Pf)* predisposing infants to severe malaria is unknown.

**Methods:** We undertook a cross-sectional study of 195 infants aged 6-24 months in Cameroon. Infants were determined to be parasitaemic by microscopy or RDT, and their disease severity classified based on WHO criteria. The EBV infection status of each child was determined using a standard serological classification system, and the magnitude, breadth, and invasion blocking capacity of the anti-*Pf* antibody response were quantified.

**Results:** 26.7% of children were serologically positive for acute EBV infection, and the highest proportion of severe malaria cases was in children with primary acute EBV. An elevated magnitude and breadth of the antibody response with increased *in vitro* invasion-blocking capacity was observed in children with acute EBV but circulating parasitaemia *in vivo* was similar.

**Conclusion:** Acute EBV infection is a risk factor for developing severe malaria in children 6-24 months. Targeting EBV infection in young children may be beneficial in protecting against the development of severe *falciparum* malaria in children living in malaria-endemic areas.

**Key points:** Acute EBV infection in infants increases the risk of severe falciparum malaria. This does not appear to be due to an EBV-induced impairment of the anti-*Plasmodium* humoral immune response which is elevated in magnitude, breadth and function.

## Introduction

Every year approximately 90% of the ∼250 million clinical cases of malaria are confined to Sub-Saharan Africa resulting in over 600,000 deaths with children under the age of 5 and pregnant women being the most affected. *Plasmodium falciparum (Pf)*, the most virulent malaria parasite species, is responsible for most of the infections and fatalities, attributed to untreated severe syndromes such as such as severe malarial anemia (SMA), acute respiratory distress syndrome and cerebral malaria ^[1]^. Exposure to endemic malaria and primary EBV infection, acquired via exposure to maternal saliva^[2]^, both occur in early childhood. In sub-Saharan Africa, up to 97% of children have experienced primary EBV infection by 12 months of age^[3]^ after maternal antibodies have waned^[4]^.

Immunity that controls *Plasmodium* infection levels and clinical symptoms is thought to build over time with repeated infections. Immunity to severe malaria, in particular non-cerebral severe malaria, develops after just 1 or 2 *Plasmodium* infections in areas of high transmission^[5]^. Not all children develop severe disease^[6]^. It is not fully understood why some infants cannot acquire immunity to severe, lethal disease once protective maternal antibodies wane. Acute EBV infection is generally asymptomatic in young children^[7]^ but can lead to suppression of humoral immunity to unrelated antigens in young children and adults^[8, 9]^. Several studies have shown impaired anti-EBV cytotoxic T cell responses in co-infected children^[10, 11]^ and this has been associated with higher EBV viremia ^[12, 13]^ and reactivation^[14]^. Increased EBV viremia has been associated with increased malaria attacks in co-infected children in Gabon^[14]^.

The highest *Pf* parasitaemia levels are observed in children 6 - 12 months and coincide with the age range where most children in central Africa become infected with EBV^[15]^. Interactions between EBV and *Plasmodium* infections can lead to the loss of immune mechanisms preserving viral latency and the development of an EBV-related malignancy known as endemic Burkitt’s lymphoma (eBL)^[16]^. However, few details are known regarding the potential impact of primary EBV infection on the severity of *Pf* infections.

Humoral immunity protects from SMA and is correlated with survival^[17]^. Published data argue that the generation of antibodies reacting to parasite proteins like the variant antigen *Pf* erythrocyte membrane protein-1 (*Pf*EMP-1) on the iRBC surface^[18]^ are a critical mechanism of anti-malarial immunity^[19]^. Passive transfer of antibodies from adults immune to malaria can treat children with severe malaria^[19]^. Antibodies contribute to the control of blood stages by several mechanisms, including the promotion of antibody-dependent phagocytosis of iRBCs^[20]^ and blocking invasion of RBCs^[21]^. This non-sterilizing immunity to malaria is short-lived and requires constant reinfections to be maintained^[22]^. In children, short-lived antibody responses are not always boosted upon reinfection with *Plasmodium*^[23]^, suggesting that the establishment of memory may be defective. However, no mechanism has been defined. Because immunity to malaria infection is generated when EBV exposure and infection occur, understanding the role of EBV in developing anti-malarial immunity to prevent severe disease in early life is essential.

Here, we show that acute primary EBV infection increases the likelihood of developing severe malaria in 6-24-month-old children. This effect was not seen in children who were serologically negative for EBV or had serological evidence of latent EBV infection. Although this was associated with an increase in circulating parasite density, acute EBV infection appeared to increase the magnitude and breadth of the anti-*Pf* humoral response that could better block invasion of *Pf* into new RBCs, suggesting that acute EBV infection did not impair anti-*Plasmodium* humoral responses. Thus, acute EBV must interfere with another component of antimalarial immunity essential for preventing severe malaria. Nevertheless, this study shows that acute EBV infection in young infants is a risk factor for developing severe malaria and supports that an efficacious EBV vaccine may protect infants from the consequences of eBL while reducing their risk for severe malaria.

## Materials and methods

### Ethical approval

The study was approved by the Cameroon National Ethics Committee on Human Health Research (Ethical clearance No: 2016/01/683/CE/CNERSH/SP), the Emory University IRB (#IRB00084993) and the University of Utah IRB (#IRB_00099332). Administrative Research Authorization was obtained from the Ministry of Public Health, Cameroon (Number 6310716).

### Study design

This was a cross-sectional study carried out between 2015 and 2017 in four different district hospitals (Efok, Obala, Efoulan and Nkoleton) and one pediatric hospital (Centre Mère et Enfant Chantal Biya) in the central region of Cameroon, where malaria transmission is perennial and hyperendemic with *Pf* responsible for over 99% of malaria cases^[24]^. This study included children aged ≤ 24 months who presented at the Emergency Units for consultation with at least one symptom of malaria as diagnosed by the clinical officer. Not all children with fever were found to be positive for *P. falciparum* and were classified as malaria negative with fever attributed to other causes. Children with evidence or history of meningitis, encephalitis, a history of developmental delay, or other neurological conditions were excluded. The demographics of the study population are shown in **Table 1**.

**Table 1:**
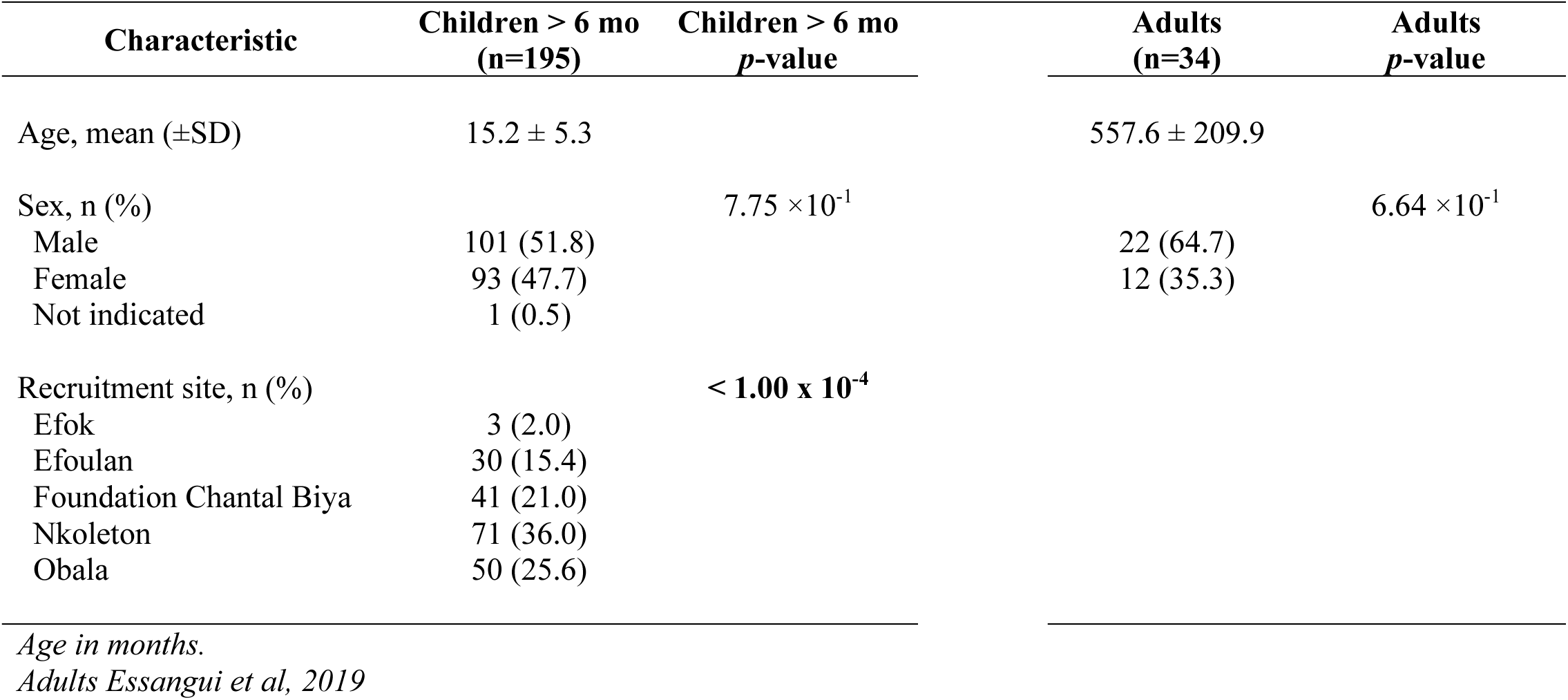
Cohort characterization of Epstein Barr Virus on Malaria infection in young children.

*Plasmodium* infections were diagnosed by Rapid Diagnostic Test (RDT) and microscopy according to standard WHO procedures. Severe malaria was defined according to the WHO criteria. Uncomplicated malaria was defined as the presence of fever or history of fever in the last 48 hours with *Plasmodium*-positive thick blood smears, but without signs of severe malaria. *Plasmodium-* infected children (severe or uncomplicated) were treated according to national guidelines. Subjects negative both on thick smears and rapid diagnostic tests were used as uninfected controls. Samples from adults over 20 years of age living in Esse Health District, Centre Region Cameroon^[25]^ were used as positive controls for studying the acquired immune response.

Venous blood samples were collected in EDTA tubes before anti-malaria treatment and transported in refrigerated boxes to the Molecular Parasitology Laboratory of Centre Pasteur Cameroon. Plasma was obtained by centrifugation at 3,000 RPM for 5 min and immediately stored at −80 °C.

### Anti-*Pf* ELISA

Plasma antibody levels were determined by ELISA using *Pf* protein soluble extracts as previously described^[25]^. Total soluble antigens were extracted from the *Pf* 3D7 following freeze-thaw fractionation procedures^[25]^.

### Epstein-Barr Virus diagnostic ELISAs

ELISAs to EBV viral capsid antigen (VCA) and EBV nuclear antigen (EBNA)-1 were quantified using commercial ELISA kits according to the manufacturer’s recommendations (GenWay Biotech). The applied cut-off was 10 Units / ml for all three ELISAs (VCA IgM, VCA IgG and EBNA-1 IgG). Infection status (Negative, acute, or latent infection) was defined as previously described^[26]^ (**Table 2).**

**Table 2:**
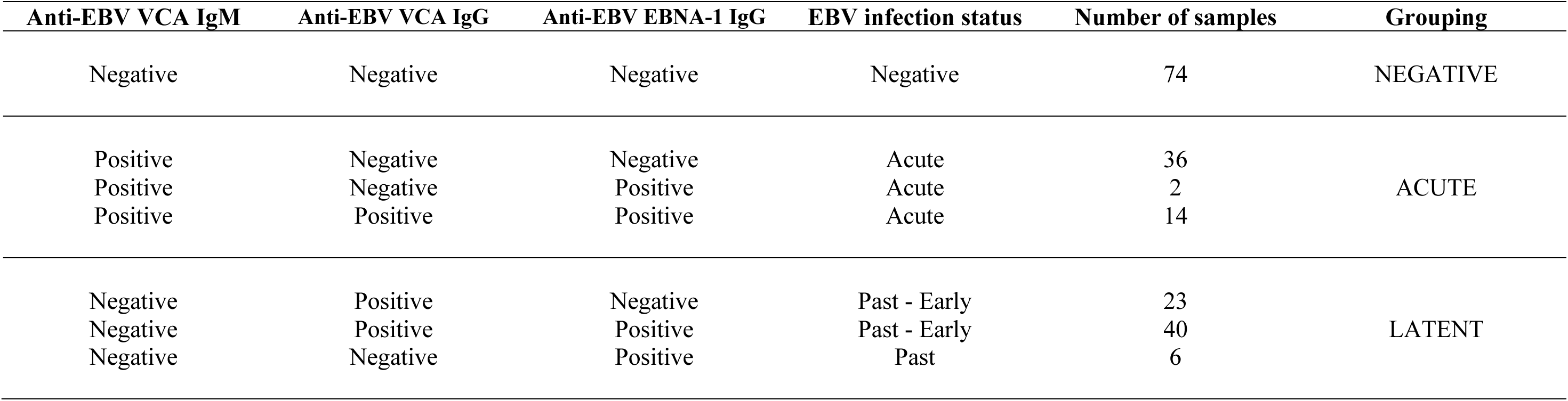
Classification of Epstein-Barr Virus status in young children based on patients’ serological profiles.

### *Pf* protein microarray analysis

All sequences for this array were derived from a single reference genome (3D7). Microarrays (Antigen Discovery Inc., Irvine, CA) were printed and probed following previously published methods^[27]^. Plasma samples were diluted 1:200 in Protein Array Blocking buffer (GVS Life Sciences, Sanford, ME) supplemented with 10% *Escherichia coli* lysate (Antigen Discovery Inc., Irvine, CA) to block any reactivity against *E. coli* in the arrayed *in vitro* transcribed/translated (IVTT) proteins. Bound IgG and IgM were then visualized using appropriate secondary antibodies. Slides were scanned on a GenePix 4200AL (Molecular Devices, Sunnyvale, CA) and spot intensities quantified using ScanArray Express software (Perkin Elmer, Waltham, MA). The pixel intensity for each antigen was corrected by subtracting background based on the average of ‘control spots’ containing the components of IVTT reactions lacking plasmid template. The minimum pixel intensity value+1 was then added to the entire dataset to ensure all intensities were positive. These intensities were used to calculate the estimated Area Under the Curve (eAUC) using the trapezoidal method for each sample. Mean intensities were considered a proxy for the titre of IgG and IgM present against *Pf* antigen, and the eAUC was used as a proxy for the total IgG and IgM against all *Pf* antigens in the array. The seropositivity rate for each sample of antigens was determined by using the corrected intensity data to calculate the fold-over IVTT control (FOC). FOC normalization provides a relative measure of the specific antibody binding to the nonspecific antibody binding to the IVTT controls. Antigens with an FOC value of greater than 2 were considered seropositive for IgG and IgM.

### Growth inhibition assay

Asexual stages of *Pf* NF54 (Patient E isolate) was cultured using standard methods. Trophozoites and early schizonts were magnetically enriched and resuspended to 1% parasitaemia in human blood. The samples were transferred to 96 well-round bottom plates (Greiner Bio-One), and the hematocrit was adjusted to 5% using media containing plasma from the available children samples at a 1:200 dilution. Cytochalasin D at 10 µM and malaria naïve serum from individuals in the United States were used as positive and negative controls, respectively. The initial parasitaemia at plating and after invasion about 24 hours after plating were quantified using flow cytometry by staining cultures with 10 µg/mL Hoechst 33342 before acquisition. Percent inhibition was calculated by subtracting background events from an uninfected sample and dividing the percentage of infected erythrocytes with normal human serum from the negative control. This number was then converted to a percentage and subtracted from 100 to determine the growth inhibitory capacity for each sample.

### Statistical analyses

Data were Log_10_, or Arcsine transformed to improve model fit for statistical analyses. Contingency tables followed by chi squared tests were used to assess the relationship between EBV status and malaria infectious status. To evaluate the impact of EBV status on antibody responses to *Pf* infection; a one-way analysis of variance (ANOVA), followed by a fisher least significant difference (LSD) *post hoc* test was used to determine statistical significance between groups. Quantile regressions were performed to estimate the effect of malaria infection status on *Pf* antibody responses within EBV groups. A generalized regression was used to assess the efficacy of plasma samples on parasite invasion blocking. Comparisons were considered significant when LSD or FDR-adjusted *p*-values for multiple hypothesis correction were < 0.05. Statistical tests were performed with JMP 17 Pro.

## Results

### Children with acute EBV infection are more likely to have severe malaria

Participants were grouped based on serological responses to viral capsid antigen (VCA) and the latency-associated protein EBNA-1^[26]^. Children negative for EBV were negative for both antigens. A serological response to either VCA or EBNA-1 was classified as EBV-exposed. In this study acute primary EBV infection was defined as a positive serological response to VCA but not EBNA −1. Latent infection was defined as lacking an IgM response to VCA and/or IgG response to EBNA-1. Within latent infection we identified individuals who also had a response to VCA but not EBNA-1 (early latent or lack of conversion to latent) or positive for IgM VCA, IgG VCA and IgG EBNA-1 which may have resulted from reactivation of a latent infection. These classifications are summarized in **Table 2**.

In the cohort of individuals 62.1% were serologically positive for EBV infection with 26.7% having an acute EBV infection (**Fig. 1A**, **Table 3**).The breakdown of EBV categorization according to malaria status is shown in **Table 3**. Similar to mouse models of acute gammaherpesvirus / *Plasmodium* co-infections^[28]^, the presence of acute EBV infection was associated with severe malaria in our cohort (**Fig. 1B**). Although children who were serologically negative for EBV tended to be younger (**Fig. 1C**), the circulating parasitaemia in microscopically positive individuals did not differ from older children who had latent EBV infection (**Fig. 1D**). However acute EBV infection was associated with significantly higher circulating *Pf* parasitaemia compared to EBV negative or latent individuals, and as expected adults had lower parasitaemia than infants(**Fig. 1D**).

**Figure 1.**
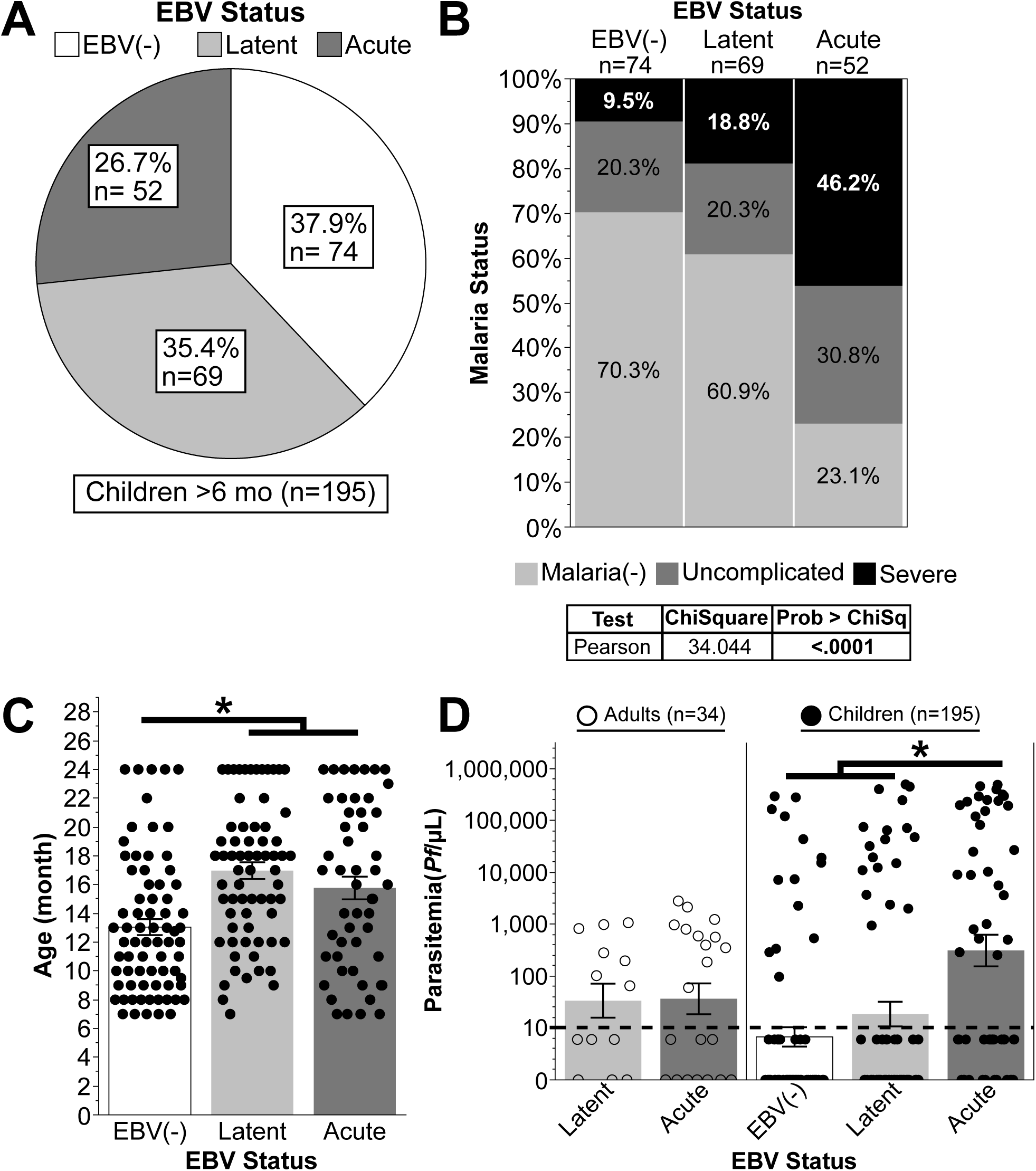
Acute EBV infection is associated with a greater probability of a severe malaria episode in infants <24 months old. (A) Distribution of EBV status groupings in young children 6-24 months of age. (B) Mosaic plot of malaria status within EBV group. (C) Distribution of ages within EBV status. (D) Distribution of circulating *Pf* iRBCs in children grouped by EBV status. Children were positive for malaria if *Pf* infection was detected by either RDT or microscopy. Parasitaemia values for RDT positive but microscopy negative children were considered below the limit of detection and given an aribitrary value of ‘5’. Adults served as positive controls for all experiments. Error bars represent SEM. * *p*<0.05 by ANOVA with LSD pairwise comparisons.

**Table 3:**
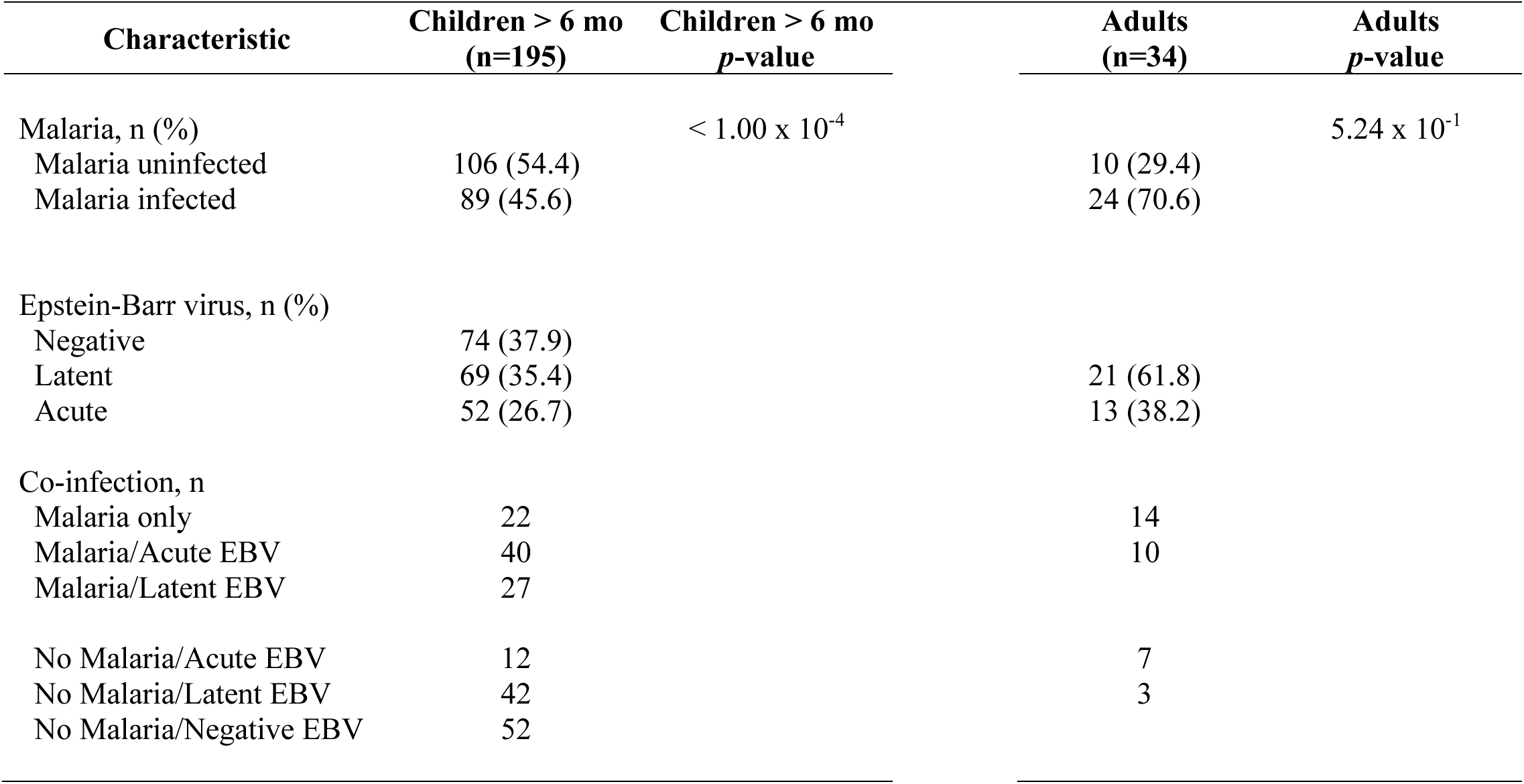
Cohort characterization of Epstein Barr Virus on Malaria infection in young children.

| Characteristic | Children > 6 mo<br>(n=195) | Children > 6 mo<br>*p*-value | Adults<br>(n=34) | Adults<br>*p*-value |
| --- | --- | --- | --- | --- |
| Malaria, n (%) |  | < 1.00 x 10<sup>-4</sup> |  | 5.24 x 10<sup>-1</sup> |
| Malaria uninfected | 106 (54.4) |  | 10 (29.4) |  |
| Malaria infected | 89 (45.6) |  | 24 (70.6) |  |
| Epstein-Barr virus, n (%) |  |  |  |  |
| Negative | 74 (37.9) |  |  |  |
| Latent | 69 (35.4) |  | 21 (61.8) |  |
| Acute | 52 (26.7) |  | 13 (38.2) |  |
| Co-infection, n |  |  |  |  |
| Malaria only | 22 |  | 14 |  |
| Malaria/Acute EBV | 40 |  | 10 |  |
| Malaria/Latent EBV | 27 |  |  |  |
| No Malaria/Acute EBV | 12 |  | 7 |  |
| No Malaria/Latent EBV | 42 |  | 3 |  |
| No Malaria/Negative EBV | 52 |  |  |  |

### Children with acute EBV infection had the highest anti-*Pf* response antibody response

A previous study in a mouse model of gammaherpesvirus – *Plasmodium* infection^[28]^ turned a non-lethal mouse model of *Plasmodium* infection into a lethal infection due to suppression of humoral responses. Thus, we hypothesized that children with acute EBV infection would have an inferior antibody response to *Pf* antigens. Contrary to the hypothesis, the antibody responses against *Pf* were increased in children with acute EBV (**Fig. 2A**). Because the ELISA measured IgM, IgG, and IgA simultaneously against lysates, the samples were then probed against protein microarrays, which enabled the assessment of the magnitude and breadth of the anti-*Pf* response for both IgG and IgM. Similar to the ELISA, children with acute EBV infection had a greater magnitude of *P. falciparum*- reactive IgM (**Fig. 2B**) and IgG (**Fig. 2C**). The breadth of the IgM (**Fig. 2D**) and IgG (**Fig. 2E**) response was also greater in children with acute EBV infection. As expected, adults had the highest measurements compared to children (**Figs. 2A-E**).

**Figure 2.**
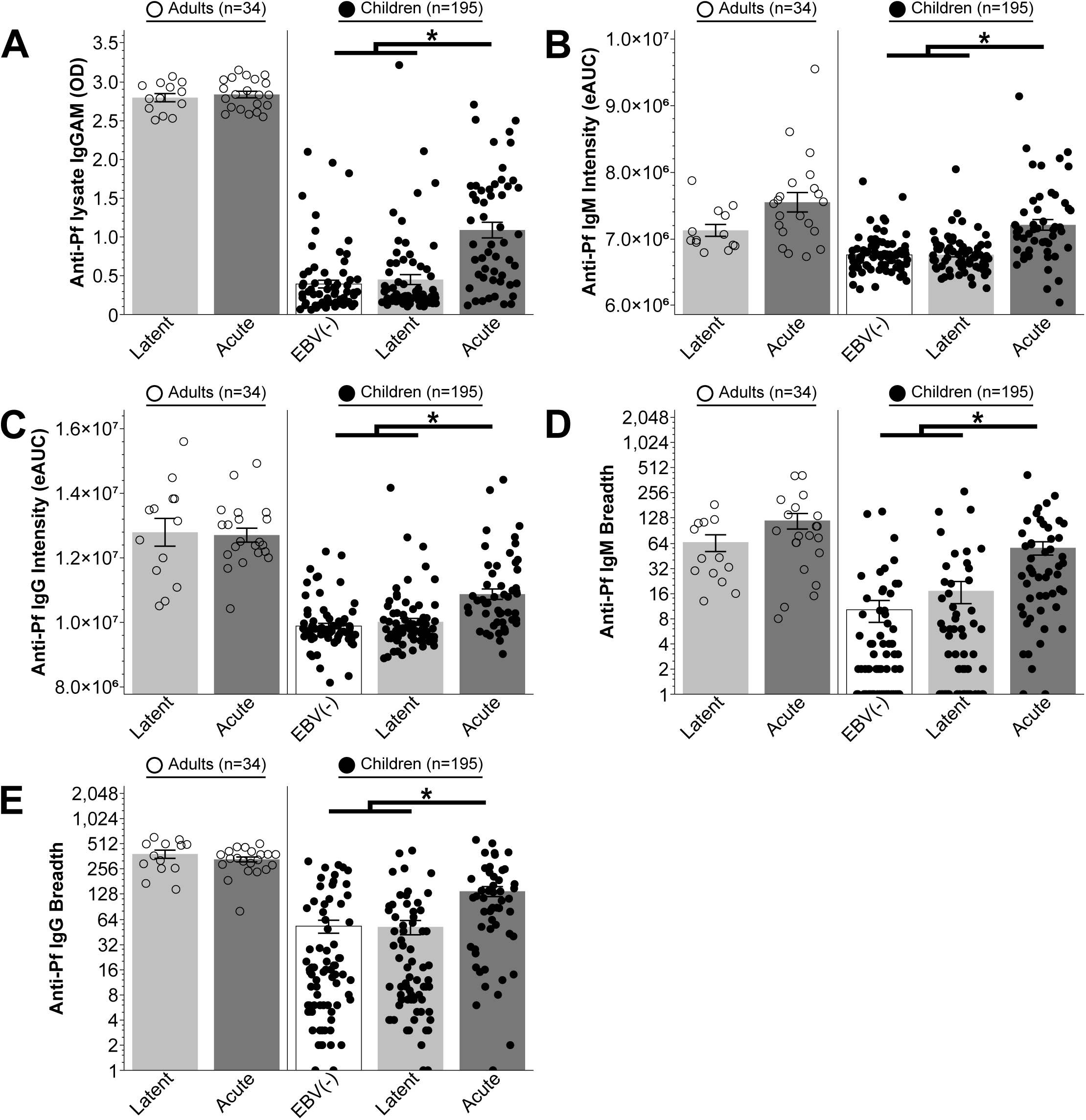
Infants with acute EBV infection had more anti-*Pf* reactive antibodies relative to those with latent EBV or those who had no serological evidence of EBV infection. (A) ELISA measurement of total anti-*Pf* IgG in plasma from adults or children between EBV status. (B) Comparison of IgM intensity estimated by area under the curve (eAUC) between EBV status (C) Comparison of IgG intensity estimated by area under the curve (eAUC) between EBV status. (D) Comparison of IgM breadth between EBV status. (E) Comparison of IgG breadth between EBV status. Adults served as positive controls for all experiments. Error bars represent SEM. * p<0.05 by ANOVA with LDS pairwise comparisons.

### The breadth of the anti-*Pf* response is associated with EBV infection independently of age

When the EBV status is subsetted by malaria status and severity, EBV infection was associated with increased magnitude of the antibody response against *Pf* lysates (**Fig 3A**) and breadth of both IgM and IgG responses, regardless of whether the infection was latent or acute (**Figs. 3B** and **3C**, **Table 4**). Whilst this may in part due to the older age of EBV-infected children (**Fig. 1C**), age did not contribute to anti-*Pf* humoral responses measured within the cohort of this study (**Table 4**). Thus, the intensity and breadth of the anti-*Plasmodium* reactive humoral response was driven by *Plasmodium* infections in both EBV latent and acute infections.

**Figure 3.**
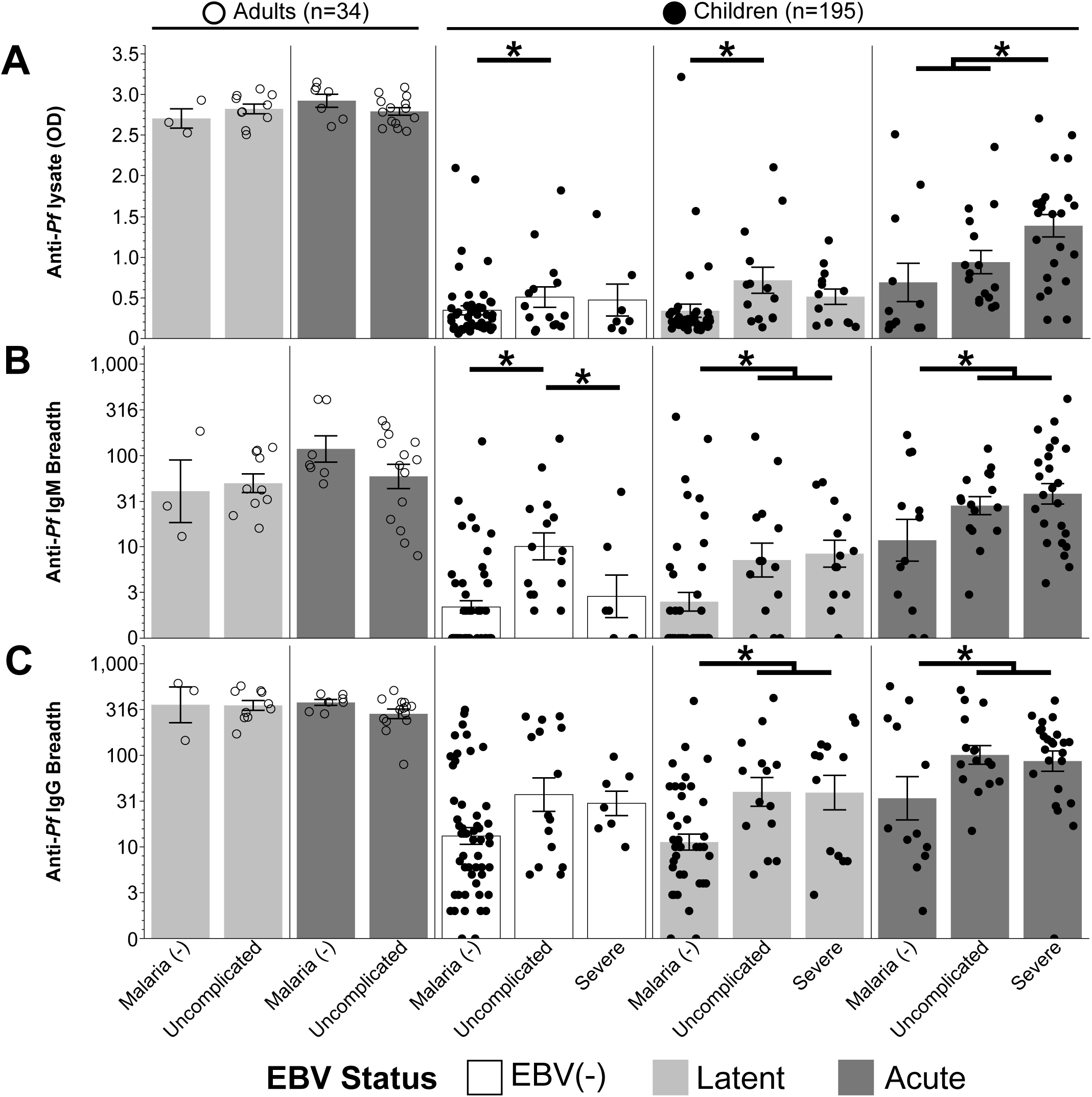
Symptoms of malaria are associated with greater humoral anti-*Pf* responses in EBV-infected infants. (A) ELISA measurement of total anti-*Pf* IgG in plasma from adults or children between malaria status within EBV groups. (B) Comparison of IgM breadth between malaria status within EBV groups (C). Comparison of IgG breadth between malaria status within EBV groups. Adults served as positive controls for all experiments. * p<0.05 by quantile regression with LDS or FDR-adjusted p-values for multiple hypothesis correction. Error bars represent SEM.

**Table 4:** Effects tests of anti-*Pf* antibody responses with epidemiological characteristics in young children with Epstein Barr Virus.

| Effects Test | Sum of Squares | F Ratio | Prob > F |
| --- | --- | --- | --- |
| **IgM Intensity** |  |  |  |
| Age (month) | $3.04 \times 10^{-4}$ | $6.58 \times 10^{-1}$ | $4.19 \times 10^{-1}$ |
| Weight | $4.33 \times 10^{-4}$ | $9.35 \times 10^{-1}$ | $3.35 \times 10^{-1}$ |
| Hb (g/dL) | $2.79 \times 10^{-6}$ | $6.00 \times 10^{-3}$ | $9.38 \times 10^{-1}$ |
| HCT (v/v %) | $4.25 \times 10^{-5}$ | $9.18 \times 10^{-2}$ | $7.62 \times 10^{-1}$ |
| Body Temp (C°) | $1.50 \times 10^{-4}$ | $3.24 \times 10^{-1}$ | $5.70 \times 10^{-1}$ |
| EBV Status | $1.03 \times 10^{-2}$ | 11.12 | **$< 1.00 \times 10^{-4}$** |
| **IgM Breadth** |  |  |  |
| Age (month) | $5.62 \times 10^{-1}$ | 1.96 | $1.64 \times 10^{-1}$ |
| Weight | $5.22 \times 10^{-2}$ | $1.82 \times 10^{-1}$ | $6.70 \times 10^{-1}$ |
| Hb (g/dL) | $5.35 \times 10^{-4}$ | $1.90 \times 10^{-3}$ | $9.66 \times 10^{-1}$ |
| HCT (v/v %) | $5.57 \times 10^{-2}$ | $1.94 \times 10^{-1}$ | $6.60 \times 10^{-1}$ |
| Body Temp (C°) | $1.18 \times 10^{-1}$ | $4.12 \times 10^{-1}$ | $5.22 \times 10^{-1}$ |
| EBV Status | 10.61 | 18.48 | **$< 1.00 \times 10^{-4}$** |
| **IgG Intensity** |  |  |  |
| Age (month) | $1.507 \times 10^{-4}$ | $1.34 \times 10^{-1}$ | $7.15 \times 10^{-1}$ |
| Weight | $1.64 \times 10^{-4}$ | $1.46 \times 10^{-1}$ | $7.03 \times 10^{-1}$ |
| Hb (g/dL) | $1.09 \times 10^{-3}$ | $9.68 \times 10^{-1}$ | $3.27 \times 10^{-1}$ |
| HCT (v/v %) | $3.07 \times 10^{-2}$ | 1.01 | $4.58 \times 10^{-1}$ |
| Body Temp (C°) | $2.74 \times 10^{-3}$ | 2.43 | $1.21 \times 10^{-1}$ |
| EBV Status | $8.77 \times 10^{-3}$ | 3.90 | **$2.26 \times 10^{-2}$** |
| **IgG Intensity** |  |  |  |
| Age (month) | $9.58 \times 10^{-1}$ | 2.64 | $1.06 \times 10^{-1}$ |
| Weight | $2.51 \times 10^{-1}$ | $6.93 \times 10^{-1}$ | $4.07 \times 10^{-1}$ |
| Hb (g/dL) | $5.79 \times 10^{-2}$ | $1.59 \times 10^{-1}$ | $6.90 \times 10^{-1}$ |
| HCT (v/v %) | $2.04 \times 10^{-2}$ | $5.63 \times 10^{-2}$ | $8.13 \times 10^{-1}$ |
| Body Temp (C°) | $8.28 \times 10^{-1}$ | 2.28 | $1.33 \times 10^{-1}$ |
| EBV Status | 2.99 | 4.13 | **$1.79 \times 10^{-2}$** |

### Plasma from parasitaemic infants with acute EBV exhibits better invasion blocking capacity

Since antibody magnitude and breadth do not necessarily predict neutralization, a growth inhibition assay was performed with samples where enough plasma was available. The greater magnitude and breadth of humoral response in children with acute EBV infection was associated with a better invasion inhibition capacity *in vitro* and was similar to adults (**Fig. 4A**). This was specific to *Pf* infection because individuals with acute EBV infection but were RDT and microscopically negative for *Pf* did not inhibit parasite growth *in vitro* (**Fig. 4B**). Surprisingly, the best correlate of invasion blocking capacity was IgM magnitude and breadth (**Table 5**), particularly in children who were positive for *Plasmodium* infection (**Fig 4B**).

**Figure 4.**
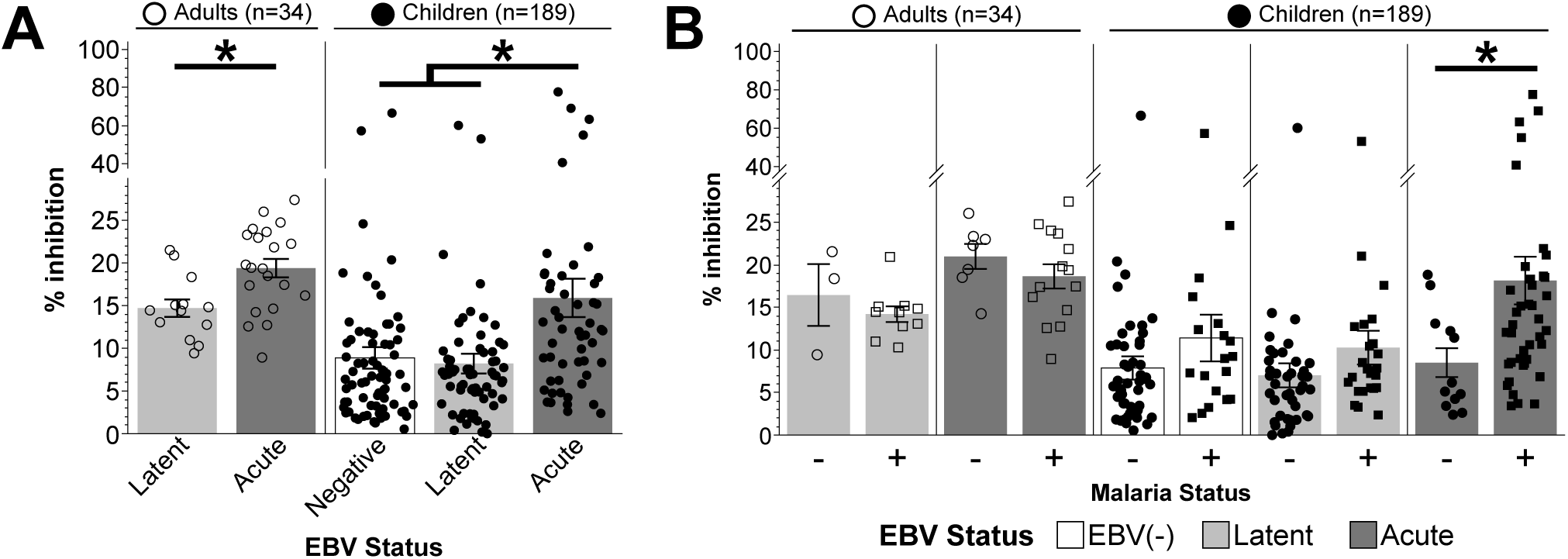
Acute EBV infection is associated with greater *in vitro* invasion inhibition in children positive for *Pf* infection. (A*) Pf* NF54 growth inhibition by plasma between EBV status. Data was averaged between two experimental replicates. Error bars represent SEM. A One-way ANOVA with the Fishers LSD post hoc test was carried out and *p* < 0.05 was considered significant. (B) Comparison of malaria status within EBV groups. Adults served as positive controls for all experiments. \**p* < 0.05 by a generalized regression with an FDR-adjusted *post hoc* test. Adults served as positive controls for all experiments.

**Table 5:** Correlation of *Pf* invasion blocking with antibody responses in young children with Acute Epstein Barr Virus.

| Infection Status | Spearman's $\rho$ | Prob>\|$\rho$\| | FDR Adj P value |
| --- | --- | --- | --- |
| **Acute EBV** |  |  |  |
| IgM Intensity | $5.44 \times 10^{-1}$ | **< $1.00 \times 10^{-4}$** | **$1.00 \times 10^{-4}$** |
| IgM Breadth | $4.74 \times 10^{-1}$ | $4.00 \times 10^{-4}$ | **$8.00 \times 10^{-4}$** |
| IgG Intensity | $1.52 \times 10^{-1}$ | $2.87 \times 10^{-1}$ | $3.82 \times 10^{-1}$ |
| IgG Breadth | $1.07 \times 10^{-1}$ | $4.50 \times 10^{-1}$ | $4.50 \times 10^{-1}$ |
| **Malaria Status** |  |  |  |
| Positive (+) |  |  |  |
| IgM Breadth | $3.96 \times 10^{-1}$ | $1.14 \times 10^{-2}$ | **$3.05 \times 10^{-2}$** |
| IgG Breadth | $-3.62 \times 10^{-1}$ | $2.16 \times 10^{-2}$ | **$4.33 \times 10^{-2}$** |
| Negative (-) |  |  |  |
| IgM Breadth | $4.45 \times 10^{-1}$ | $1.47 \times 10^{-1}$ | $1.97 \times 10^{-1}$ |
| IgG Breadth | $7.06 \times 10^{-1}$ | $1.02 \times 10^{-2}$ | **$3.05 \times 10^{-2}$** |

## Discussion

The overlap in endemicity between *Pf* infections and EBV in children is associated with the development of eBL^[16]^, a non-aggressive non-Hodgkin lymphoma that manifests in 5-9 year old children in sub-Saharan Africa^[29]^. However, the effect of acute EBV infection on *Pf* infections has not been reported despite the possibility of immunological interactions when young children are coinfected^[30]^. As shown in a mouse model of gammaherpesvirus / *Plasmodium* infection^[28]^, the severity of malaria can be increased in infants with an acute EBV infection.

The hypothesis that EBV may suppress antibody responses to an incoming infection rests on the fact that EBV largely resides in germinal center B cells^[31]^. Although there has been some evidence suggesting that *P. vivax* responses are impaired in a study involving 56 adults living in a *P. vivax* endemic area of the Amazon, there has never been a thorough analysis undertaken when infants first become infected with EBV^[32]^. Unlike in mouse MHV68-*P. yoelii* XNL infections^[28]^ the mechanism does not appear to be due to EBV-induced suppression of anti*-Pf* humoral responses. In contrast, our data suggests that EBV infection augments anti-*Pf* humoral responses, in particular the magnitude and breadth of the IgM response, which can exert substantial invasion-blocking capacity *in vitro*. Despite an augmented IgM response, circulating parasitaemia was higher in infants presenting with acute EBV infection.

There are several possible explanations underlying this finding. First, an augmented IgM response correlating with increased parasitaemia may be an effect rather than a cause of EBV infection. The IgM induced may be cross-reactive with peptides present on the *Pf* array and induced as part of the polyclonal activation of IgM B cells by EBV^[33]^. Secondly, EBV infection may induce suppression of immune mechanisms other than antibody production that may also contribute to control of *Pf* iRBCs. These could include modulation of innate responses^[34]^. For example, EBV is known to inhibit interferon regulatory factors (IRFs) to prevent induction of type 1 interferons^[35]^. Although type 1 interferons have been shown in mouse models to suppress priming of Tfh by B cells resulting in reduced IgM and IgG responses^[36]^ type 1 interferon is also needed to augment CD4 T cell-derived IFN-γ^[37]^ which activates several parasite clearance mechanisms but is also correlated with pathogenesis^[38]^.

Additionally, EBV transcribes a homolog of IL-10^[39]^ that likely serves to increase B cell proliferation and expand the latency reservoir but can suppress Th1 responses. IL-10 helps protect against the pathogenesis of malaria but may impair control of blood stage parasitaemia. The serum concentration of IL-10 during infection with *Pf* is associated with higher parasite densities and less effective parasite clearance in Tanzanian children^[40]^.

This study departs from the current thinking that the main consequence of infant primary EBV infection is simply the eventual development of eBL in some children and suggests it is a risk factor for the development of severe malaria. The outcome of this work may extend to other childhood infections prevalent in sub-Saharan Africa, particularly related herpesviruses such as Karposi’s sarcoma-associated herpes virus and cytomegalovirus, but further studies are required.

## Data Availability

All data produced in the present study are available upon reasonable request to the authors

## Acknowledgements

We extend our gratitude to the Department of Pediatrics and the medical staff at Obala, Efoulan, and Nkoleton District Hospitals, as well as the Chantal Biya Foundation Hospital, for assistance with diagnostics and phlebotomy. We are also thank the clinical staff, patients, and their guardians at these institutions for their participation in this study. Finally we thank the field staff and, most importantly, residents of the Esse Health district in Cameroon who participated in this study. The following reagent was obtained through BEI Resources, NIAID, NIH: *Plasmodium falciparum*, Strain NF54 (Patient Line E), MRA-1000, contributed by Megan G. Dowler.

## Author contributions

WTG performed the growth inhibition assays and data analysis; BF, BU, SK, EE, CD, and CN performed field sample and serological analysis of the processed samples; AJ, HD and PF performed the antibody array; SHS devised the study and reviewed the paper; SN, CE and PK supervised the study; CJJ analysed the data, supervised WTG, and wrote the paper; LSA devised the study, supervised the study, analyzed data and wrote the manuscript; TJL devised the study, analyzed the data and wrote the manuscript

